# The World Smells Different in Parkinson’s Disease

**DOI:** 10.64898/2026.02.22.26346819

**Authors:** Michal M. Andelman-Gur, Sagit Shushan, Kobi Snitz, Gal Pinchasof, Danielle Honigstein, Lior Gorodisky, Aharon Ravia, Adi Ezra, Neomi Hezi, Tanya Gurevich, Noam Sobel

## Abstract

Olfactory decline is a well-established aspect of Parkinson’s disease (PD) and is considered one of its earliest signs, often preceding motor symptoms by years to decades. However, because olfactory impairment is also common in healthy aging and other medical conditions, current olfactory tests that score performance (odor detection, discrimination, and identification) lack disease specificity. In contrast to performance scores, olfactory perceptual fingerprints are derived from odor ratings and sniffing behavior, and provide a stable measure of how the world smells to an individual. To test the hypothesis that olfactory perceptual fingerprints may provide a disease-specific marker, we obtained them in three cohorts: Individuals with PD (n=33), healthy age-matched controls (n=33), and critically, in participants with non-PD olfactory dysfunction (n=28). Consistent with previous results, a standard clinical olfactory test detected impairment in both PD and non-PD olfactory dysfunction, but failed to distinguish between these two groups. In contrast, olfactory perceptual fingerprints detected impairment, and distinguished PD from non-PD olfactory dysfunction at 88% accuracy (SVM classification, leave-one-out cross validation, 90% sensitivity, 85% specificity, *P*=3.2×10^-4^), or 94% accuracy after matching age and sex (SVM classification, leave-one-out cross-validation, 100% sensitivity, 88% specificity, *P*=0.0047). The difference between PD related and unrelated olfactory decline was particularly evident in sniffing behavior: Whereas both healthy participants and non-PD olfactory decline groups decreased sniff duration in response to unpleasant odors (−12.5% and −11.36% respectively), individuals with PD paradoxically increased sniff duration (+1.69%; P=4.5×10^-5^). Thus, PD was marked not by loss of olfactory performance, but by a distinct shift in olfactory perception. These findings imply that olfactory perceptual fingerprints provide for a disease-specific marker in PD.

## Introduction

An olfactory impairment in PD was first identified 50 years ago, in a study that found elevated detection thresholds (i.e., poorer performance) for the odorant amyl acetate (banana) in 10 of 22 relatively young individuals with PD (*1*). The advent of more sensitive clinical olfactory tests starting with the University of Pennsylvania Smell Identification Test (UPSIT) (*2*), and later the Sniffin’ Sticks battery (*3*), now imply olfactory impairment in well over 90% of individuals with PD (*4, 5*). Moreover, the olfactory impairment in PD can precede motor symptoms by years or even decades (*4, 6, 7*). The reason for this olfactory impairment in PD is attributed to specific disease-related atrophy in the neural substrates of olfaction. There is indeed remarkable overlap between the early brain pathophysiology of PD and the olfactory system, at both the cellular (*8–10*) and structural (*4, 11, 12*) levels. Whereas it is widely accepted that this brain damage underlies the olfactory impairment in PD, the cascade that drives it remains unknown. One alternative is that the disease process begins somewhere in the brain but then selectively targets olfactory brain structures (*4*). An alternative hypothesis is that the disease may follow infection by some unknown pathogen, perhaps prion-like, that may enter the brain through the nose, and therefore initially act along its olfactory path of entry (*12, 13*).

Regardless of its origins, the olfactory impairment in PD has become a widely used supporting PD biomarker, albeit with one significant limitation: it is non-specific. Olfactory impairments are common, especially after the COVID19 pandemic, where infection can drive long-lasting olfactory loss (*14*). Moreover, olfactory impairment is evident across neurodegenerative diseases (*15, 16*), and is in fact common in “healthy aging” (*17*). Even within PD itself, the severity of olfactory impairment varies, particularly between *LRRK2*-associated cases and idiopathic PD (*18*). Therefore, olfactory testing has provided for only supporting evidence in the PD diagnostic routine. Notably, olfactory testing may gain specificity in combination with MRI (*19, 20*), but dependence on brain imaging limits access. An alternative to performance-based testing is perceptual characterization. Olfactory perceptual fingerprints are a tool that characterizes how the world smells to an individual (*21*). Fingerprints are generated from descriptor ratings applied to odorants. The fact that different individuals may use descriptors differently (*22*) does not affect these fingerprints, because they are constructed within an individual, reflecting how a given set of odorants relate to each other. These fingerprints remain stable over time (*21*), and reflect unique olfactory perceptual phenotypes (*23–26*). Moreover, how the world smells to an individual can further be estimated by how they sniff it. Sniffing is odorant, and particularly odorant-intensity and valence specific (*27, 28*). Thus, by characterizing the olfactory sniff-response we obtain an added non-verbal measure of perception. As PD is now recognized to involve multisensory perceptual alterations – spanning visual, auditory, tactile, and temporal perception and implicating putative basal-ganglia networks (*29–33*), we posited that our olfactory measures might reflect this broader perceptual shift in PD.

Here we combine these measures to ask whether they can provide for an olfactory measure with specificity for PD. We apply these tests in three cohorts: non-demented (mostly Stage II) individuals with PD, non-PD-related olfactory loss, and healthy age-matched controls. We assured heterogeneity in the non-PD-related olfactory loss group. This was important because had all the non-PD-related olfactory loss been for one reason, e.g., post-COVID, we would not be able to determine whether we are detecting PD or detecting post-COVID. Also, we verified that the age-matched controls exhibited olfactory performance typical for their age group (i.e., age-appropriate mild-hyposmia) to distinguish PD from age-typical olfactory decline (*34*). We find that perceptual characterization allows for accurate classification of PD at 89% accuracy vs. healthy controls, and critically, at 88% accuracy vs. diverse non-PD-related olfactory loss. This level of accuracy increases to 94% after matching the groups for sex and age. Thus, perceptual characterization provide for an olfactory test specific for Parkinson’s disease.

## Results

### Impaired olfactory performance is non-specific for Parkinson’s disease

All participants completed the Sniffin’ Sticks test, a standard clinical olfactory assessment that measures olfactory detection threshold, olfactory discrimination, and olfactory identification (*34*). Given the abnormal distribution of scores (Kolmogorov-Smirnov test, all *D*>0.78, all *P*<4.2×10^-17^), we proceeded to analyze them using a nonparametric approach with Bonferroni correction for multiple comparisons. We observed significant differences between groups (Kruskal-Wallis Test, Total score: c*^2^*(_2_)=48.2, *P*=3.4×10^-11^, Fig. 1d; Threshold: c*^2^*_(2)_=44.3, *P*=2.4×10^-10^, Fig. 1a; Discrimination: c*^2^*_(2)_=24.2, P=5.6×10-6, Fig. 1b; Identification: c*^2^*_(2)_= 38.4, *P*=4.4×10-9, Fig. 1c), demonstrating that in comparison to healthy controls, both the PD and OD groups performed poorly on the Sniffin’ Sticks test. This was evident both in total score (Post-hoc Wilcoxon rank-sum test with Bonferroni correction, median±SD: Control=30.5±5.5, PD=16.5±5.9, OD=12.4±7.2, Control vs. PD: *Z*=6.1, *P*=1.2×10^-8^, Cliff’s δ=0.88, 95% Confidence Interval (CI)=0.72-0.95; Control vs. OD: *Z*=5.6, *P*=2.8×10^-7^,Cliff’s δ=0.83, 95% CI=0.64-0.93; PD vs. OD: *Z*=1.9, *P*=NS, Cliff’s δ=0.3, 95% CI=-0.03–0.6; Fig. 1d) and on each subpart of the test (Threshold: Control=7.5±2.7, PD=2.7±2.4, OD=1±1.4, Control vs. PD: *Z*=4.6, *P*=5.4×10^-5^, Cliff’s δ=0.65, 95% CI=0.41-0.8; Control vs. OD: *Z*=6.2, *P*=7.9×10^-9^, Cliff’s δ=0.9, 95% CI=0.78 - 0.96; PD vs. OD: *Z*=2.7, *P*=NS, Cliff’s δ=0.4, 95% CI=0.1–0.64; Fig. 1a. Discrimination: Control=11±2.7, PD=8±2.6, OD=6±2.9, Control vs. PD: *Z*=3.9, *P*=9.4×10^-4^, Cliff’s δ=0.57, 95% CI=0.31-0.75; Control vs. OD: *Z*=4.3, *P*=1.8×10^-4^, Cliff’s δ=0.64, 95% CI=038-0.81; PD vs. OD: *Z*=1.13, *P*=NS, Cliff’s δ=0.17, 95% CI=-0.13–0.4; Fig. 1b. Identification: Control=13±2.0, PD=5±2.7, OD=5±4.1, Control vs. PD: *Z*=6.2, *P*=5.9×10^-9^, Cliff’s δ=0.88, 95% CI=0.7-0.96, Control vs. OD: *Z*=4.3, *P*=2.2×10^-4^, Cliff’s δ=0.63, 95% CI=0.31-0.82; PD vs. OD: *Z*=0.2, P=NS, Cliff’s δ=0.03, 95% CI=-0.3–0.3; Fig. 1c). In other words, consistent with previous studies (*4, 35*), the clinical test was sensitive for detecting olfactory loss, but was non-specific as to its cause. Both PD and OD groups performed equally worse than controls, and no different from each other.

**Figure 1:**
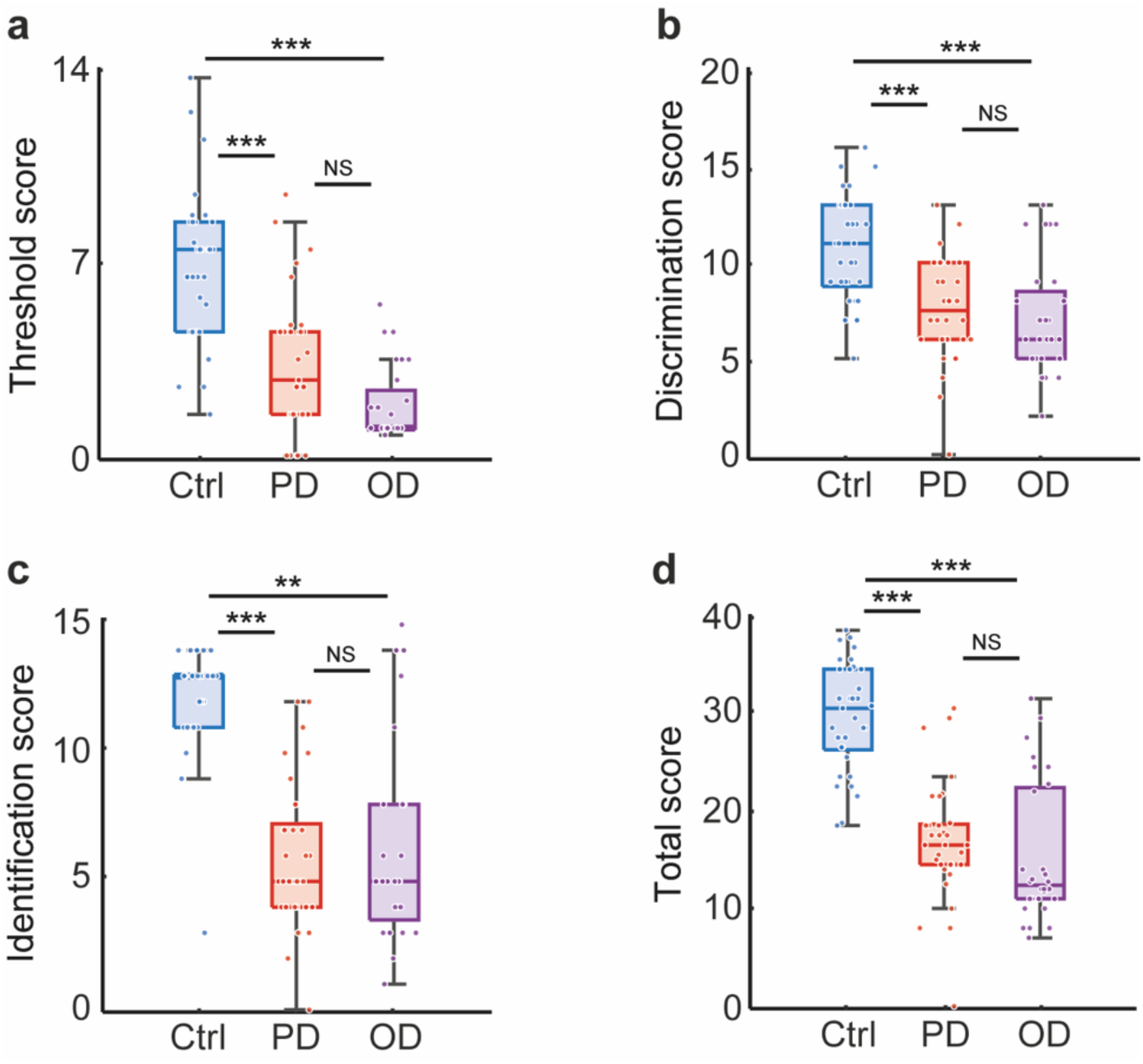
Impaired olfactory performance is non-specific for Parkinson’s disease. **a-d.** Box plots illustrating Sniffin’ Sticks scores for the PD group (red, n = 33), OD group (purple, n = 28), and healthy controls (blue, n = 33), in the three different subparts of the test: Threshold (a), Discrimination (b), Identification (c), and in the Total score (d). Central lines indicate median, top and bottom edges of the box indicate 75th and 25th percentiles, respectively. Whiskers reflect 1.5 times the interquartile range. Each circle represents a participant. *P £ 0.05, **P £ 0.01, ***P £ 0.001.

Whereas the PD and OD groups were matched by olfactory performance, they were slightly but significantly mismatched for age and sex (OD: mean age=58.1±11.5; 13 women; PD: mean age: 64.2±7.5; 4 women). To ask whether these differences accounted for a portion of our results, we ran a linear regression on olfactory scores. More specifically, we tested whether age and sex in the OD and PD groups predicted total score in the clinical test using linear regression. Assumptions were checked via residual plots and a Q-Q plot. The model explained R^2^=0.06 (adjusted R^2^=0.012) of the variance (F_[3,56]_=1.2, *P*=0.3). Age (*β* =-0.147, SE=0.09, *t*=-1.6, *P*=0.11), and sex were not significant (*β* =0.8, *t*=0.4, *P*=0.7) (Supplementary Fig. 1). In addition, if we combine the PD and OD groups, we observed no sex difference in performance of the Total Sniffin’ Sticks score (mean women=14.9±6.8), mean men=16.68±6.6, *t*-Welch=-0.89, *P*=0.38, Supplementary Fig. 1). These results imply that the slight differences in age (mean age 65.8 vs. 58.1) and sex (number of women 4 vs. 13) between the PD and OD groups did not underlie olfactory performance differences in these cohorts. After controlling for sex and age, in the PD group, medications had no detectable effect on the Sniffin’ Sticks Total score (ANCOVA multiple linear regression, model R²=0.12, *P*=0.41). In the OD group, etiology had modest influence on Total Sniffin’ Sticks score (model R²=0.38, *P*=0.033). Idiopathic OD presented with better performance relative to the grand mean (*β*=+6.01±2.04, *P*=0.0076), whereas other etiologies were non-significant in their effect on olfactory performance.

### The world smells different in Parkinson’s disease

Intensity and pleasantness are the primary dimensions of olfactory perception (*36, 37*). There is evidence that both of these perceptual dimensions are indeed altered in PD (*38–40*), but whether they serve to discriminate PD from OD is unknown. To ask whether our cohorts differ on these dimensions, we asked participants to rate the perceived intensity and pleasantness of three different stimuli presented in sniff-jars: 1. “Pleasant”: a suprathreshold concentration of the odorant citral that smells like lemon and is typically considered pleasant. 2. “Unpleasant”: a suprathreshold concentration of a mixture of asafetida and skatole that smells fecal and is typically considered unpleasant. 3. “Blank”: an empty jar. Ratings were made on a visual-analogue scale (VAS) ranging from 1 to 100 (Supplementary Fig. 2,3). Odorants were deliberately presented at high, robustly suprathreshold levels to maximize detection in participants with impaired olfaction.

Given the abnormal distribution of the ratings (Kolmogorov-Smirnov test, all *D*>0.84, all *P*<0.001), we proceeded to analyze them using a nonparametric approach with Bonferroni correction for multiple comparisons. When comparing absolute ratings, we observed a significant difference in perceived intensity across groups in both odorants, but not in the blank condition (Kruskal-Wallis test, intensity, pleasant odorant: c*^2^*_(2)_=25.8, *P*=2.5×10^-6^; unpleasant odorant: c*^2^*_(2)_=7.7, *P*=0.02; Blank: c*^2^*_(2)_=2.76, *P*=NS, Supplementary Fig. 3). This stemmed from reduced perceived intensity for the pleasant odorant only in the OD group (Post-hoc Wilcoxon rank-sum test with Bonferroni correction, median±SD, Pleasant: Control=78±13.5, PD=78±22.6, OD=16±33.1; Unpleasant: Control=89.5±19.8, PD=81±28.7, OD=50±41.4; Blank: Control=4±6.8, PD=3±13.6, OD=3±19.9, Pleasant: Control vs. PD: *Z*=0.7, *P*=NS, Cliff’s δ=0.1, 95% CI=-0.19-0.39, Control vs. OD: Z=4.8, *P*=1.7×10^-5^, Cliff’s δ=0.7, 95% CI=0.48-0.86, PD vs. OD: Z=4.05, P=4.5×10^-4^, Cliff’s δ=0.6, 95% CI=0.36-0.8; Unpleasant: Control vs. PD: Z=1.9, P=NS, Cliff’s δ=0.3, 95% CI=-0.004-0.54, Control vs. OD: *Z*=2.6, *P*=NS, Cliff’s δ=0.4, 95% CI=0.07-0.64, PD vs OD: *Z*=1.6, *P*=NS, Cliff’s δ = 0.26, 95% CI=-0.07-0.5; Blank: Control vs. PD: *Z*=0.2, *P*=NS, Cliff’s δ=0.02, 95% CI=-0.26-0.3, Control vs. OD: *Z*=1.5, *P*=NS, Cliff’s δ=0.2, 95% CI=-0.08-0.5, PD vs OD: *Z*=1.3, *P*=NS, Cliff’s δ=0.2, 95% CI=-0.1-0.5, Supplementary Fig. 3). The groups did not differ from each other in their intensity ratings for the unpleasant odorant and for the blank (Unpleasant: all *Z*<2.6, all *P>*0.08, Blank: all *Z*<1.5, all *P*>0.99). Whereas the above compared absolute ratings, one can postulate a dulling of perception without loss of relative properties. To assess this, we repeated the above analysis, this time comparing ratings relative to blank within each participant. This reanalysis (Supplementary Fig. 2), as well as reapplying this initial question to data obtained later in this study (Supplementary Fig. 4), generated the same general outcome. In sum, only the OD group had reduced perceived intensity for the pleasant odorant.

Whereas in perceived intensity individuals with PD were like controls, and differed from OD, this was not the case for perceived pleasantness. When comparing absolute ratings, the groups significantly differed in pleasantness ratings (Kruskal-Wallis test, pleasantness, pleasant odorant: c*^2^*_(2)_=23.0, *P*=1.0×10^-5^, unpleasant odorant: c*^2^*_(2)_=14.6, *P*=0.0007, Blank: c*^2^*_(2)_=1.8, *P*=NS, Supplementary Fig. 3). This reflected emergence of the expected pleasantness ratings in controls, but less so in PD and OD (Post-hoc Wilcoxon rank-sum test with Bonferroni correction, median±SD, Pleasant: Control=81.5±15.9, PD=50±26.2, OD=50±17.6; Unpleasant: Control=7±14.8, PD=28±24.9, OD=39±23.0; Blank: Control=50±14.0, PD=50±12.5, OD=50±12.1; Pleasant: Control vs. PD: *Z*=3.3, *P*=0.006, Cliff’s δ=0.5, 95% CI=0.21-0.71, Control vs. OD: *Z*=5.0, *P*=3.9×10^-6^, Cliff’s δ=0.7, 95% CI=0.54-0.88, PD vs OD: Z=1.0, *P*=NS, Cliff’s δ=0.15, 95% CI=-0.15-0.43; Unpleasant: Control vs. PD: Z=-3.9, *P*=6.7×10^-4^, Cliff’s δ=-0.6, 95% CI=-0.76-(−0.33), Control vs. OD: Z=-3.3, *P*=0.009, Cliff’s δ=-0.5, 95% CI=-0.71-(−0.19), PD vs OD: Z=0.3, *P*=NS, Cliff’s δ=0.04, 95% CI=-0.26-0.34; Blank: Control vs. PD: *Z*=1.06, *P*=NS, Cliff’s δ=0.13, 95% CI=-0.11-0.36, Control vs. OD: *Z*=0.67, *P*=NS, Cliff’s δ=0.08, 95% CI=-0.14-0.3, PD vs OD: *Z*=-0.6, *P*=NS, Cliff’s δ=-0.07, 95% CI=-0.3-0.17, Supplementary Fig. 3). Again, repeating this analysis using ratings relative to blank in each participant (Supplementary Fig. 2), as well as reapplying this question to data obtained later in this study (Supplementary Fig. 4), generated the same general outcome.

Taken together, individuals with PD are like controls in their perceived intensity estimates, but like OD in their perceived pleasantness estimates. This dissociation fits a shift in perceptual mapping rather than generalized sensory loss, echoing non-olfactory perceptual alterations reported in PD (*30*). To ask whether these patterns of intensity and pleasantness perception could serve to classify PD, we applied an SVM classifier to the ratings and used Bayesian optimization to automatically select the optimal SVM configuration for classification (*41*). We then tested results using leave-one-out cross validation, such that testing was on participants who were not in the learning set. Finally, to assess the significance of results we conducted a bootstrap analysis where we randomly shuffled the labels of participants, and reconducted the classification 10,000 times. We found that we could classify PD from healthy controls at 77% accuracy (AUC=0.79, 84% specificity: 27 of 32 healthy, 69% sensitivity: 20 of 29 PD, Bootstrapped *Z*=2.71, *P*=0.003, Supplementary Fig. 3) and from OD at 71% accuracy (AUC=0.74, 56% specificity: 15 of 27 OD, 86% sensitivity: 25 of 29 PD, Bootstrapped *Z*=2.68, *P*=0.003, Supplementary Fig. 3). We conclude that perceptual ratings of intensity and pleasantness alone can provide for modest but significant group classification.

Whereas intensity and pleasantness reflect primary dimensions of olfactory perception, more detailed perceptual ratings allow derivation of individually specific olfactory perceptual fingerprints (*21*). To ask whether such fingerprints may uncover a perceptual shift in PD, we asked participants to rate 10 different monomolecules that span physio-chemical olfactory space (Supplementary Fig. 4,5) along 11 stable descriptors (*42*), using VAS scales from 1 to 100 (see Methods). We reiterate that subjective use of language has no impact on these fingerprints. For example, “spicy” may indeed mean one thing to participant A and a different thing to participant B. But the fingerprint calculates the difference in “spiciness” across odors within each participant, thus, the only important thing is that a given participant uses the same notion of “spiciness” across the odorants they rate. In other words, perceptual fingerprints use a subjective construct to generate an objective odor space (*21*). We projected these fingerprints into PCA space and observed that individuals with PD tend to inhabit a restricted subspace of perception (Fig. 2a). To initially quantify this observation, we first asked whether there is greater perceptual similarity within groups vs. between groups. In other words, we asked whether the olfactory perceptual world of a PD patient is more like that of other individuals with PD than it is like OD patients or controls.

**Figure 2:**
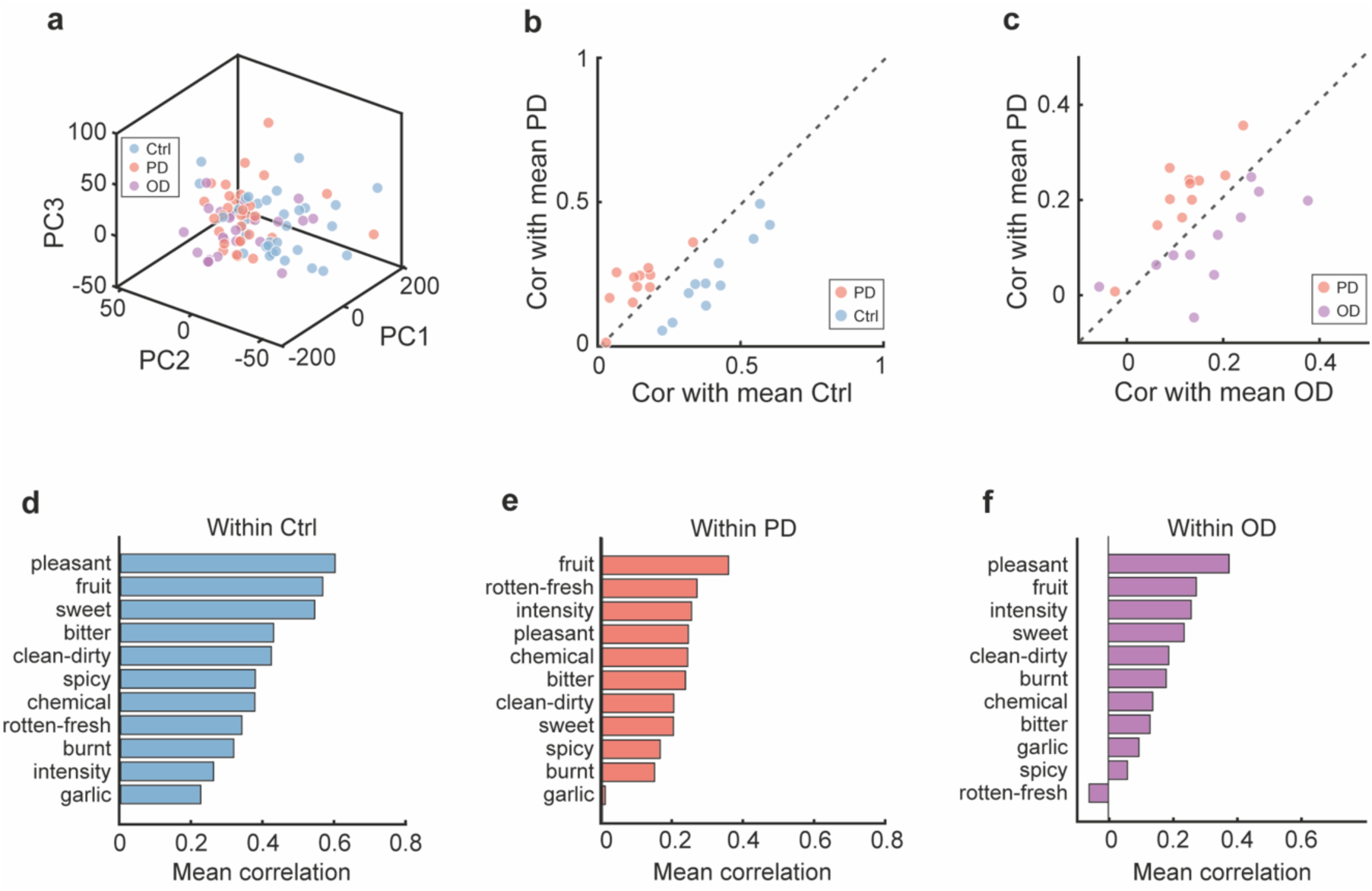
The world smells different in Parkinson’s disease. **a**, Principal Component Analysis (PCA) for olfactory fingerprints for all groups (Ctrl: blue, n=33, PD: red, n=33, OD: purple, n=28). b-c, Scatter plots comparing correlation coefficients for each descriptor in each group: healthy controls and participants with PD (b), individuals with OD and with PD (c). Each color circle represents a different descriptor (mean correlation). d-f, Mean correlation per each descriptor within each group. Descriptor names are indicated.

For each individual, we calculated the mean rating along each descriptor for each odorant. We omitted the specific participant’s rating from their group mean. Then, we calculated the correlation (using Pearson’s correlation) of each individual with the mean of their group and the other groups. We found that individuals with PD were significantly more correlated with mean PD perception than they were with mean normosmic control perception (Wilcoxon signed rank test, mean±SD, Control: r=0.4±0.12 vs. 0.24±0.14, *W*=0, *P*=9.7×10^-4^; PD: r=0.21±0.09 vs. 0.14±0.08, *W*=2, *P*=0.003, Fig. 2b) or with mean OD perception (Wilcoxon signed rank test, mean±SD, PD: r=0.21±0.09 vs. 0.12±0.07, *W*=0, *P*=9.7×10^-4^; OD: r=0.17±0.12 vs. 0.1±0.09, *W*=9, *P*=0.03, Fig. 2c). To further quantify specificity, we computed similarity matrices between ratings of odorants per group (Fig. 2g-i). To do so, we calculated Spearman’s correlation between mean odorant ratings in each group. Then, we tested whether the correlation matrices are significantly different from each other using RV coefficient and evaluated *P*-values using 10,000 bootstrapped permutations (Bonferroni corrected for multiple comparisons). We found that the groups were significantly different from one another (Control vs. PD: RV coefficient=0.8, *P*=0.02, Control vs. OD: RV coefficient=0.85, *P*=0.01, PD vs. OD: RV coefficient=0.93, *P*=0.01), suggesting that the pattern of correlations between odorants differ significantly between groups. To assess internal consistency, we repeated the analysis on random half-split within each group: the split-halves were similar in Control and PD (pattern reproduced), but diverged in OD (split-halves differed) (Figure 3a-i). In other words, there is an olfactory perceptual framework that is typical of PD.

**Figure 3.**
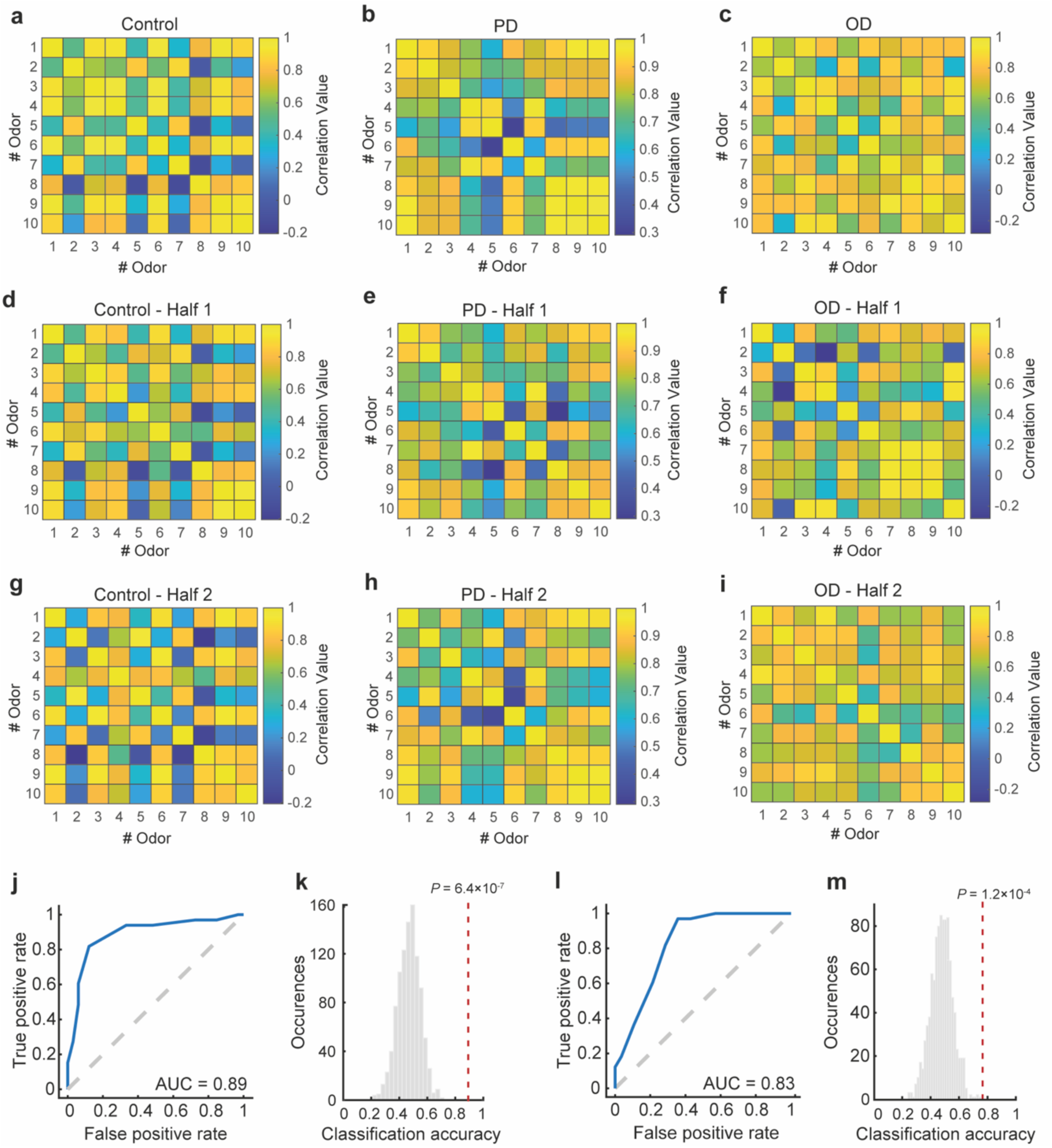
**a-c**, Similarity matrices between mean odorant ratings in each group, computed by Spearman’s correlation. For each group, similarity matrix between mean odorant ratings was computed by each half of the group, for control (d,g), PD (e,h) and OD (f,i). Odorant numbers and correlation values are indicated. j-m, ROC curves for detecting PD from healthy controls (j) and for detecting PD from OD participants (l) using the majority vote per descriptor logistic regression classifier. Histogram of classification accuracy using logistic regression for shuffled labels data of PD and control (k) and PD and OD (l) with actual classification accuracy marked by red dashed line. P-values are marked on top of the line.

We next asked whether adding these ratings to the intensity and pleasantness ratings could improve PD classification. For each individual, we separated ratings by descriptor (e.g., pleasantness, intensity), resulting in 11 distinct rating types. We then ran the classifier to predict a classification (“PD,” “Control,” or “OD”) for each descriptor, yielding 11 predictions per individual. A majority vote was applied across all descriptors to determine the final classification based on the proportion of votes (*43*). For example, if a participant was classified as PD for six descriptors and as Control for five, their final classification would be PD with a score of 6/11 (≈0.545). Using this approach, we classified PD from controls at 85% classification accuracy (56 of 66, bootstrapped *Z*=4.8, *P*=6.4×10^-7^, Fig. 3j-k), with 88% sensitivity (29 of 33) and 82% specificity (27 of 33), and PD from OD at 77% accuracy (47 of 61, bootstrapped *Z*=3.6, *P*=1.2×10^-4^, Fig. 3l-m), with 82% sensitivity (27 of 33) and 71% specificity (20 of 28). In other words, classification using all the perceptual descriptors is significant, but not overwhelmingly better than using intensity and pleasantness alone. In sum, the world smells different to individuals with PD than it does to both OD patients and controls, and this difference can provide for significant classification.

### Odor sniffing is different in Parkinson’s disease

Precise measurements of nasal airflow provide an objective non-verbal measure of perception as a function of the sniff response. The sniff response is a fast sensory-motor feedback loop where sniff magnitude is inversely proportional to odorant intensity and valence, can be driven by sensory input, cognition, or both (*44*) and its neural circuitry is dissociable from the brainstem mechanisms that pace involuntary respiration, recruiting an olfactomotor loop spanning olfactory cortex, basal ganglia, and respiratory effectors (*28, 45*). Of the various sniff parameters, sniff duration is particularly sensitive to odor properties (*46*). We therefore compared normalized sniff duration (NDU) of the first nasal inhale after presentation of odorants (pleasant, unpleasant and blank, see Methods, Fig. 4a,b). Due to the abnormal distribution of the data (Kolmogorov-Smirnov test, *P*<0.001 for all data sets), we used non-parametric tests with Bonferroni correction for multiple comparisons. At the group level, we observed that only healthy controls had significantly different sniff duration for different odorants (Kruskal-Wallis test, c*^2^*_(2)_=9.51, *P*=0.008). Within groups, both controls and OD sniffed longer for pleasant vs. unpleasant odorants, and only PD did not show this difference (Wilcoxon signed-rank test, median±SD, Control: 1.12±0.3 NDU vs. 0.98±0.26 NDU, *Z*=4.4, *P*=1.0×10^-4^, Cliff’s δ=0.29, 95% CI=0.15-0.42; OD: 1.32±0.33 NDU vs. 1.17±0.28 NDU, Z=4.3, *P*=1.7×10^-4^, Cliff’s δ=0.21, 95% CI=0.12-0.3; PD: 1.18±0.31 NDU vs 1.2±0.33 NDU, *Z*=-1.5, *P*=NS, Cliff’s δ=-0.002, 95% CI=-0.16-0.16, Fig. 4c-f). These group differences clearly emerge in the delta sniff duration that revealed how most individuals with PD in fact behaved opposite of controls, prolonging their sniff for the unpleasant odorant (Fig. 4g-h, Supplementary Fig. 5, median delta±SD, Control (−0.12±0.2 NDU) vs. PD (0.02±0.2 NDU), *Z*=-4.3, *P*=4.3×10^-5^, Cliff’s δ=-0.65, 95% CI=-0.82-(−0.36); OD (−0.09±0.2 NDU) vs. PD (0.02±0.2), *Z*=4.3, *P*=4.5×10^-5^, Cliff’s δ=0.67, 95% CI=0.39-0.84). Notably, the level of deviation from expectation in sniff duration was not correlated with disease motor severity, as assessed by the UPDRS-MDS Part III test (Spearman’s r=-0.17, *P*=NS) (the only sniff parameter correlated with disease motor severity was volume for the blank stimulus Spearman’s r=-0.44, *P*=0.016). Finally, only the control group maintained a sniff difference also for blank vs. unpleasant (Wilcoxon signed-rank test, median±SD, 1.14±0.35 NDU vs. 0.98±0.26 NDU, *Z*=4.5, *P*=5.4×10^-5^, Cliff’s δ=0.43, 95% CI=-0.16-0.6, Fig. 4e), and none of the groups sniffed with significantly different duration for the pleasant vs. blank odorants.

**Figure 4:**
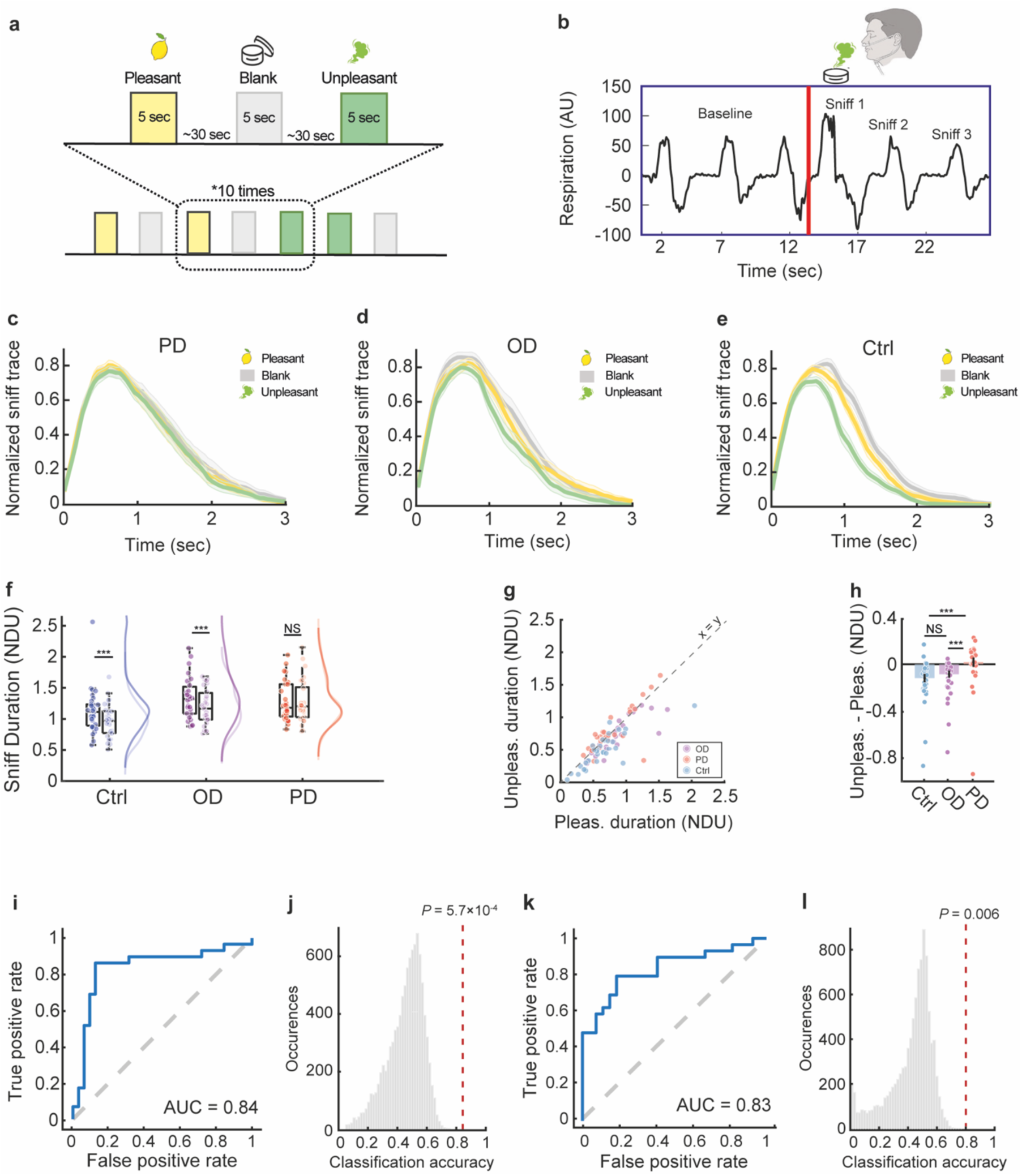
Odor sniffing is different in Parkinson’s disease. **a**, Illustration of the sniff response paradigm (*44*). b, Illustration of experimental set up. c-e, Sniff response group traces. Each trace was normalized within participant to their maximal peak value. Normalized sniff traces (mean ± s.d.) for pleasant (yellow), unpleasant (green), and blank (gray) in c. PD (n=29), d. OD (n=27), e. Control (n=32). f, Boxplots displaying duration difference between sniffs in response to pleasant (Ctrl: dark blue, OD: dark purple, PD: dark red) vs. unpleasant (Ctrl: light blue, OD: light purple, PD: light red) odorants in each group. Central lines indicate median, top and bottom edges of the box indicate 75th and 25th percentiles, respectively. Whiskers reflect 1.5 times the interquartile range. Half violin plots present the distribution in each group. Each circle represents a participant (blue: control, n=32, purple: OD, n=27, red: PD, n=29). g, Unit slope graph presenting sniff durations during pleasant (x-axis) vs. unpleasant (y-axis) odorant presentation. Each circle represents a participant (blue: control, n=32, purple: OD, n=27, red: PD, n=29). The diagonal line reflects the unit slope line (x = y) such that if points accumulate above the line then values are greater for unpleasant odorant, and if they accumulate under the line then values are greater for pleasant odorant. h, Bar graph illustrating the difference between pleasant and unpleasant sniff duration for each group. Each circle represents a participant (blue: control, n=32, purple: OD, n=27, red: PD, n=29). i-l, ROC curves for detecting individuals with PD from healthy controls (i) and for detecting individuals with PD from participants with OD (k) using an SVM classifier. Histogram of classification accuracy for shuffled labels data of PD and control (j) and PD and OD (l) with actual classification accuracy marked by red dashed line. P-values are marked on top of the line. *P £ 0.05, **P £ 0.01, ***P £ 0.001.

We next asked whether we could classify PD based on altered olfactory sniffing alone. We applied an SVM classifier to the differences in duration and volume of the pleasant and unpleasant odorants. We tested using leave-one-out cross validation such that testing was on participants not in the learning set. We employed a Bayesian optimization approach to automatically select the optimal SVM configuration for classification (*41*). Using this we discriminated PD from normosmic controls at 85% accuracy (52 of 61, AUC=0.84), 86% sensitivity (25 of 29) and 84% specificity (27 of 32), and PD from OD at 80% accuracy (45 of 56, AUC=0.83), 79% sensitivity (23 of 29) and 81% specificity (22 of 27) (Fig. 4i,k). To assess the significance of this result we conducted a bootstrap analysis where we randomly shuffled the labels of participants 10,000 times, and reconducted the classification. This implied high significance for both classifiers (PD vs. control classifier: 85% accuracy, bootstrapped Z = 3.25, P = 5.7×10-4; PD vs. OD classifier: 80% accuracy, bootstrapped Z = 2.46, P = 0.007, Fig. 4i-l).

### An olfactory test specific for Parkinson’s disease

We found that individuals with PD perceive odors differently and sniff them differently. Each one of these differences alone provided for a modest but significant classifier of PD. We now tested whether we could combine these to make for a test with true specificity for PD. We used a stacking approach: applying a meta-level classifier to the probability distribution of both base-level classifiers (*47*) (one for each data set). We performed this analysis only on individuals who participated in all experiments (32 controls, 29 PD, 27 OD). Probability score for SVM classifier predictions was calculated using a sigmoid function (*48*). We applied a third classifier (SVM) with a leave-one-out cross validation on the probability scores for each participant (one for each training data set) such that testing was on participants not in the learning set and reached an optimized classification result. As before, for each one of the two comparisons, we applied the Bayesian SVM optimization approach for tuning the classification parameters. This resulted in 89% accurate classification of PD from controls (53 of 61; 83% sensitivity (24 of 29), 94% specificity (30 of 32), AUC = 0.86, Bootstrapped *Z*=3.3, *P*=4.6×10^-4^, Fig. 5b-c), and 88% accurate classification of PD from OD (50 of 56; 90% sensitivity (26 of 29), 85% specificity (23 of 27), AUC = 0.9, Bootstrapped *Z*=3.4, *P*=3.2×10^-4^, Fig. 5d-e). As previously noted, the PD and OD cohorts were slightly but significantly different in age and sex. To ask whether these differences influenced our results, we replicated this analysis after first excluding participants incrementally until the groups did not differ in age and sex. This process retained 16 matched participants per group (Age: mean PD(n=16)=64.7±5.1, mean OD(n=16)=62.6±11.4, Welch’s *t*-test (unequal variances): *t*=-0.66, *P*=0.52, Cohen’s d=-0.23. Sex: number of women: PD=3/16, 18.8%, OD=6/16=37.5%, Fisher exact test: *P*=0.433, OR=2.6; Chi-square with Yates correction *χ²*_(1)_ = 0.62, *P*=0.432). Despite the lower power afforded by this truncated cohort size, our method significantly discriminated between the two groups at 94% accuracy (30 of 32, Bootstrapped *Z*=2.6, *P*=0.0047), AUC=0.97 (Fig. 5f-g), 100% sensitivity (16 of 16), and 88% specificity (14 of 16). We conclude that perceptual fingerprints provide for an olfactory test with specificity for PD. PD patients are heterogenous in olfactory performance, but share a pattern in olfactory perceptual fingerprints. We note that the combined duration of these tests is less than 30 minutes. Moreover, we note that perceptual fingerprints can be obtained using either of the two commonly applied clinical tests: the UPSIT and Sniffin’ Sticks. The only necessary modification to their application is in the questions asked, and in the added measure of nasal airflow.

**Figure 5:**
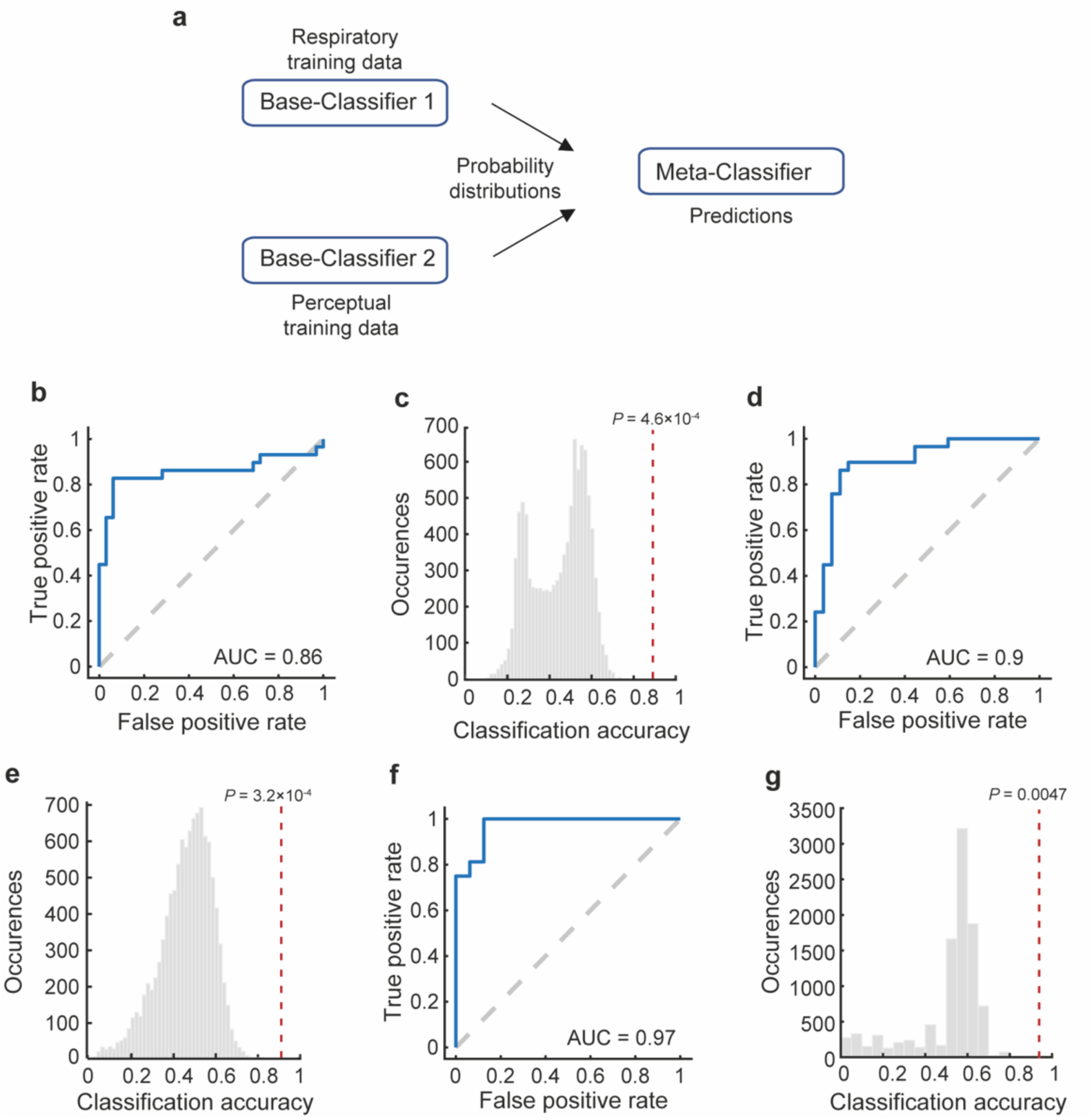
An olfactory test specific for Parkinson’s disease. **a.** Schematic design of the stacking algorithm for both data sets. b,d,f. ROC curves for detecting PD from healthy controls (b) and for detecting PD from OD participants (d) and from age- and sex-matched OD participants (f) using the stacking algorithm. c,e,g. Histogram of classification accuracy for shuffled labels data of PD and control (c), PD and OD (e) and PD and age- and sex-matched OD (g) with actual classification accuracy marked by red dashed line. P-values are marked on top of the line.

## Discussion

It is well established that there is considerable variability among PD patients in terms of the degree of their smell loss (*18*). For this reason, tests of olfactory performance per se failed to differentiate between PD and non-PD-related olfactory loss. Here we initially replicated this result, but then found that by contrast, perceptual fingerprints provided for 89% classification accuracy of PD vs. age-matched controls, and critically, 88% classification accuracy of PD vs. non-PD-related olfactory loss, with 94% accuracy after matching for sex and age. In other words, whereas PD-performance is heterogeneous, PD-perception, at least in these mostly stage-II patients, reflects a disease-specific profile. This supports a broader notion that, beyond motor symptoms (*49*), PD involves a systematic shift in perception across modalities (*30*). Critically, this classification power persisted despite variability in the non-PD related olfactory loss group. In other words, had we tested only one type of alternative olfactory loss, we would not be able to determine whether we captured a PD-profile, or the alternative profile. However, with the current heterogenous non-PD loss groups, we are more confident that we captured a genuine PD-related perceptual profile.

One may ask how the nature of the perceptual alteration we observed can be related to the brain pathophysiology of PD. In the functional hierarchy of olfaction, detection threshold is considered a very basic task (*50*), and its impairment is often taken to implicate peripheral olfactory machinery. Individuals with PD were clearly impaired at detection threshold (Fig. 1a), but unexpectedly, they were like controls in rating perceived intensity (Fig 2b-d). These two seemingly contradictory results combine to imply that the impairment in detection threshold may in fact reflect a meta-level task-related impairment that goes beyond olfaction alone. Indeed, individuals with PD are impaired at auditory detection threshold (*51*), and at most visual just-noticeable-difference (JND) tests (*52*). Thus, their impairment at olfactory detection threshold may not necessarily reflect peripheral olfactory receptor-events, and instead may reflect higher-order mechanisms related to the temporal and memory demands of any detection threshold task (unlike intensity estimates, detection threshold involves change-detection, namely a comparison across conditions separated in time). Consistent with this notion of non-peripheral involvement, the olfactory mucosa in PD remains unaffected by α-Synuclein deposition (*53*). By contrast, brain regions involved in the formation and maintenance of olfactory percepts beginning with valence and beyond are indeed sites of documented α-Synuclein deposition. This includes the olfactory bulb and anterior olfactory nucleus (*4*), ventral temporal primary olfactory cortex (*54*), and orbitofrontal secondary olfactory cortex (*55*). The perceptual shift evident here in individuals with PD, primarily reduced pleasantness of pleasant odors, is consistent with the general anhedonia in PD (*56*), and is likely associated with damage in these brain areas.

The altered olfactory perception in PD was evident not only in odorant ratings, but also in odorant sniffing. Sniffs are odorant-specific, and particularly odorant-valence-specific, in what has been termed the sniff-response (*28*). The sniff-response is preserved across species (*57, 58*), in humans it is observed in both wake (*59*) and sleep (*60*), and provides for a marker in brain pathologies ranging from autism (*61*) to disorders of consciousness (*44*), and relies on neural circuitry dissociable from the brainstem mechanisms that pace automatic respiration (*28, 45*). We had previously found that sniffing is reduced overall in PD (*62*), and here we find that sniffing is no longer odorant-specific in PD. In fact, individuals with PD took longer sniffs for the particularly unpleasant odorant we used than for the markedly pleasant odorant. In other words, they behaved opposite of both healthy controls and individuals with non-PD-related olfactory loss. Such changes might reflect a general distortion in perception-to-action coupling and sensory-motor integration, possibly arising from basal-ganglia-mediated broadening of receptive fields and imbalanced precision-weighting (*63*). Based on these results we would like to put forth the following hypothesis: The underlying assumption in the field is that PD pathology damages the neural substrates of olfaction, and this then gives rise to olfactory impairment. However, an alternative is a reversal of cause and effect: We have recently found that ongoing nasal respiration, independent of olfaction, is altered in PD (*64*). It is possible that this, together with the altered olfactory sniffing, gives rise to a form of olfactory self-deprivation in PD. Consistent with this notion of deprivation, improved breathing through aerobic training improves olfaction in PD (*65*). Olfactory deprivation is indeed a known cause of neural degradation across neural substrates with extensive overlap to the neuropathology of PD. Most notably, olfactory deprivation reduces volume of the anterior olfactory nucleus (*66*), possibly the earliest site of PD brain pathology (*67*). Olfactory deprivation also profoundly alters the neuroanatomy and neurochemistry of the olfactory bulb (*68*). Notably, the bulb is directly connected with the substantia nigra via a dopaminergic pathway (*69*). In other words, we are speculating that olfactory loss is a cause rather than a result of PD. Genetic causal association analysis suggests viability for this proposal. More specifically, whereas PD was more clearly a causal factor in hyposmia, hyposmia was nevertheless a weak yet significant causal factor in PD (*70*). Although suggesting lost olfaction as a cause rather that effect in PD is an admittedly highly speculative hypothesis, it is one that may have profound clinical implications.

Finally, our study had two limitations we would like to acknowledge: First, we did not include an added, fourth group, of non-PD neurodegeneration. Olfactory impairment is common across the neurodegenerative diseases (*15, 16*), and whether our reported specificity holds in comparison to such a non-PD neurodegeneration cohort is a necessary future step before the currently described test can become clinically useful. The second notable limitation is that we had significantly more men than women in our cohort. This stems in part from the fact than men are 1.5 times more likely than women to develop PD (*71*), but the imbalanced cohort remains a recruitment shortcoming that limits the interpretation of our results. Nonetheless, despite these two limitations, our results uncover that individuals with PD have a form of PD-typical olfactory perception, and that this can be used to classify PD not only from healthy age-matched individuals, but also from individuals with non-PD-related olfactory loss. In this, we provide an olfactory test specific to PD that could support early detection and is particularly relevant given ongoing efforts to develop disease-modifying therapies (*72–75*).

## Materials and Methods

### Participants

Between the years 2021-2024, we recruited 33 individuals with PD (mean age: 65.8 ± 7.2, 4 women, 5 left-handed), 33 matched healthy controls (mean age: 64.2 ±7.5, 4 women, 5 left-handed) and 28 OD patients (mean age: 58.1 ± 11.5, 13 women, 3 left-handed). OD patients were matched by olfactory performance to the PD group. Individuals with PD were recruited from the movement disorders unit at Tel-Aviv Sourasky Medical Center, Tel-Aviv, Israel, and OD patients were recruited from the Otolaryngology (ENT) department at Edith Wolfson Medical Center, Holon, Israel. All participants provided written informed consent to procedures approved by the ethics committees (Helsinki) of Tel-Aviv Medical Center (Protocol # 0239-20-TLV) and Wolfson Medical Center (Protocol # 140-21-WMC).

PD diagnosis was confirmed both by clinical symptoms and by ^18^F-DOPA PET/CT brain imaging, and olfactory dysfunction diagnosis was based on ENT specialist examination. To make sure the OD group would be as heterogenous as possible, patients were included based on various causes for olfactory dysfunction. Since mild hyposmia is common with aging (*34*), we a priori allowed ‘age-appropriate mild hyposmia’ in controls. The groups did not differ in cognitive performance (MoCA PD = 24.8±3.9, Control=25.7±2.0, OD=25.5±3.4, Kruskal-Wallis test, *χ^2^*_(2)_=0.39, *P*=0.8), but individuals with PD scored higher on the depression questionnaire (Beck PD=11.5±8.2, Control=4.9±4.8, OD=8.5±9.1, Kruskal-Wallis test, *χ^2^*_(2)_=14.9, *P*<0.001). Clinical and demographic characteristics are presented in Supp Table 1. All participants completed the standard olfactory tests (Sniffin’ Sticks) (*34*) and the olfactory ratings. A subset of 29 participants with PD, 32 healthy normosmic controls, and 27 OD patients completed the sniff response experiment. All individuals with PD were examined during the ON state.

### Standard Olfactory testing

All participants completed the standardized “Sniffin’ Sticks” test, consisting of threshold, discrimination, and identification subtests (*34*). Total score (TDI) ranged from 3 to 48.

### Odorants

For the olfactory perception experiment we used 10 monomolecules that span physio-chemical odor space (*42*) (Supplementary Fig. 5): Fenchone (CAS 7787-20-4, 7.6% in IPM), Isoamyl acetate (CAS 123-92-2, 25% in IPM), 3-Propylidene phthalide (CAS 17369-59-4, 7.9% in IPM), Cuminaldehyde (CAS 122-03-2, 13.6% in IPM), Strawberry glycidate 1 (CAS 77-83-8, 100%), Nonanal (CAS 124-19-6, 100%), Citral (CAS 5392-40-5, 100%), Skatole (CAS 83-34-1, 0.75% in IPM), Hexanol (CAS 111-27-3, 30% in IPM), 6-Methylquinoline (CAS 91-62-3, 10.7% in IPM); all above odorants were obtained from Sigma-Aldrich. For the sniff response experiment, we used a mixture of ‘asafetida’ (DreamAir, LLC) and skatole (CAS 83-34-1, Sigma-Aldrich) as the unpleasant stimulus, and citral (CAS 5392-40-5, Sigma-Aldrich) as the pleasant stimulus. In both experiments, the odorants (100 μL in each sample) were absorbed in a cotton pad and placed in a sniff jar. A jar with only cotton pad (without any odorant) served as the blank stimulus.

### Olfactory rating

For the olfactory perception task, participants rated 10 monomolecular odorants (see Odorants section) along 11 descriptors (Intensity, pleasantness, sweet, fruity, garlic, spicy, burnt, chemical, clean-dirty, fresh-rotten, bitter) using 100-point visual analog scales (*42*). Odorant and descriptor order were randomized; inter-trial intervals exceeded 40 seconds.

### Sniff response procedure and analysis

Nearly all participants (PD=29, Control=32, OD=27) underwent a sniff response task. Odorants (pleasant, unpleasant, or blank) were presented 10 times each in a random order (Fig. 3). Presentation duration=5 seconds, inter-stimulus-interval=∼40 seconds. Participants were cued to sniff, and nasal airflow was recorded using a miniature sensor previously described (*76*). Sniff magnitude was normalized to pre-odor baseline. Trial and sniff exclusions were based on predefined criteria that were previously published (*44*), including absent or unstable baselines and extreme outliers (>3SD than the average). Based on these criteria, 121 of 2609 trials (4.6%), 693 of 7428 baseline breaths (9.3%), and 82 of 7010 sniffs (1.2%) were excluded from the study.

### Statistical analysis

To estimate the intended number of participants we conducted a power Analysis using G*Power 3.1 (*77*) based on previous results of airflow differences in pleasant vs. unpleasant odorants in healthy participants, showing a decrease from flow pleasant = 0.918 ± 0.32 normalized flow units [NFU] to flow unpleasant = 0.665 ± 0.22 NFU (*61*) at 305 ms of sniff onset. This indicated that, at power of 0.95 and α < 0.05, we needed at least 15 participants per group.

Given non-normal data (confirmed by Kolmogorov-Smirnov test), non-parametric tests were used: Wilcoxon rank-sum or Kruskal-Wallis for group comparisons; Wilcoxon signed-rank test for within group analyses. Bonferroni correction for multiple comparisons was applied when necessary. Cliff’s δ (*78*) estimated effect sizes; Spearman correlation assessed association with disease severity; RV coefficient measured matrix similarity.

### Classification analyses

We applied support vector machine (SVM) classifiers to sniff response data and logistic regression to olfactory ratings data, both using leave-one-out cross validation. SVM parameters were optimized via Bayesian search (*41*). Analyses were conducted in JASP (version 0.17.2.1, JASP team), MATLAB (version R2023a, MathWorks, Inc.) and Python (version 3.9.12, Python Software Foundation). All raw data collected for this manuscript and code for analyses and figures are available at: https://gitlab.com/michal.andelman/world-smells-different-in-pd.

## Supporting information

Supplemental matrials

## Data Availability

All raw data collected for this manuscript and code for analyses and figures are available at: https://gitlab.com/michal.andelman/world-smells-different-in-pd.

## Acknowledgments

This study was funded by a grant from the European Research Council ERC Synergy project D2Smell awarded to NS. MAG was supported by a doctoral fellowship from the Azrieli Foundation.

## Author Contributions

Conceived idea: M.A.G., N.S.; Designed experiments: M.A.G., T.G., N.S.; Recruited participants: A.E., N.H., T.G., S.S., Programmed the sniff response experiment: D.H.; Conducted experiments: M.A.G., G.P.; Analyzed data: M.A.G., A.R., L.G., N.S.; Wrote first draft: M.A.G.; Edited final draft: All authors.

## References

1. K. Ansari, A. Johnson, Olfactory function in patients with Parkinson’s disease. Journal of chronic diseases 28, 493–497 (1975).

2. R. L. Doty, S. M. Bromley, M. B. Stern, Olfactory testing as an aid in the diagnosis of Parkinson’s disease: development of optimal discrimination criteria. Neurodegeneration 4, 93–97 (1995).

3. B. Herting, S. Schulze, H. Reichmann, A. Haehner, T. Hummel, A longitudinal study of olfactory function in patients with idiopathic Parkinson’s disease. Journal of neurology 255, 367–370 (2008).

4. R. L. Doty, Olfactory dysfunction in Parkinson disease. Nature Reviews Neurology 8, 329–339 (2012).

5. C. Murphy et al., Prevalence of olfactory impairment in older adults. Jama 288, 2307–2312 (2002).

6. A. Haehner et al., Olfactory loss may be a first sign of idiopathic Parkinson’s disease. Movement disorders 22, 839–842 (2007).

7. C. Murphy, in Sensory Science and Chronic Diseases: Clinical Implications and Disease Management. (Springer, 2022), pp. 145–158.

8. R. Pearce, C. Hawkes, S. Daniel, The anterior olfactory nucleus in Parkinson’s disease. Movement disorders: official journal of the Movement Disorder Society 10, 283–287 (1995).

9. I. Ubeda-Bañon et al., The human olfactory system in two proteinopathies: Alzheimer’s and Parkinson’s diseases. Translational neurodegeneration 9, 1–20 (2020).

10. T. J. Stevenson et al., α-synuclein inclusions are abundant in non-neuronal cells in the anterior olfactory nucleus of the Parkinson’s disease olfactory bulb. Scientific reports 10, 6682 (2020).

11. E. Wattendorf et al., Olfactory impairment predicts brain atrophy in Parkinson’s disease. Journal of Neuroscience 29, 15410–15413 (2009).

12. I. Ubeda-Bañon, D. Saiz-Sanchez, C. de la Rosa-Prieto, A. Martinez-Marcos, α-Synuclein in the olfactory system in Parkinson’s disease: role of neural connections on spreading pathology. Brain Structure and Function 219, 1513–1526 (2014).

13. C. Hawkes, B. Shephard, S. Daniel, Is Parkinson’s disease a primary olfactory disorder? Qjm 92, 473–480 (1999).

14. B. Blomberg et al., Long COVID in a prospective cohort of home-isolated patients. Nature medicine 27, 1607–1613 (2021).

15. R. I. Mesholam, P. J. Moberg, R. N. Mahr, R. L. Doty, Olfaction in neurodegenerative disease: a meta-analysis of olfactory functioning in Alzheimer’s and Parkinson’s diseases. Archives of neurology 55, 84–90 (1998).

16. C. Marin et al., Olfactory dysfunction in neurodegenerative diseases. Current allergy and asthma reports 18, 1–19 (2018).

17. N. E. Rawson, Olfactory loss in aging. Science of Aging Knowledge Environment 2006, pe6–pe6 (2006).

18. L. Silveira-Moriyama et al., Olfactory heterogeneity in LRRK2 related Parkinsonism. Movement disorders 25, 2879–2883 (2010).

19. J. Mei, C. Tremblay, N. Stikov, C. Desrosiers, J. Frasnelli, in Medical Imaging 2021: Computer-Aided Diagnosis. (SPIE, 2021), vol. 11597, pp. 320–327.

20. S. Brosse, C. Tremblay, I. Mérida, J. Frasnelli, Specific structural changes in Parkinson’s disease-related olfactory dysfunction compared to others forms of olfactory dysfunction. Frontiers in Neural Circuits 18, 1503841 (2024).

21. L. Secundo et al., Individual olfactory perception reveals meaningful nonolfactory genetic information. Proceedings of the National Academy of Sciences 112, 8750–8755 (2015).

22. A. Majid, Cultural factors shape olfactory language. Trends in cognitive sciences 19, 629–630 (2015).

23. T. Weiss et al., Human olfaction without apparent olfactory bulbs. Neuron 105, 35–45. e35 (2020).

24. K. Snitz et al., An olfactory self-test effectively screens for COVID-19. Communications medicine 2, 34 (2022).

25. E. Drnovsek, A. Haehner, M. Bensafi, T. Hummel, Olfactory perceptual fingerprints of people with olfactory dysfunction and healthy controls. Laryngoscope Investigative Otolaryngology 9, e1267 (2024).

26. E. Drnovsek et al., An olfactory perceptual fingerprint in people with olfactory dysfunction due to COVID-19. Chemical Senses 48, bjad050 (2023).

27. M. Bensafi, S. Pouliot, N. Sobel, Odorant-specific patterns of sniffing during imagery distinguish ‘bad’and ‘good’olfactory imagers. Chemical senses 30, 521–529 (2005).

28. B. N. Johnson, J. D. Mainland, N. Sobel, Rapid olfactory processing implicates subcortical control of an olfactomotor system. Journal of neurophysiology 90, 1084–1094 (2003).

29. L. Avanzino, M. Fiorio, A. Conte, Actual and illusory perception in Parkinson’s disease and dystonia: a narrative review. Frontiers in neurology 9, 584 (2018).

30. F. Permezel, J. Alty, I. H. Harding, D. Thyagarajan, Brain networks involved in sensory perception in Parkinson’s disease: a scoping review. Brain Sciences 13, 1552 (2023).

31. C. Ding et al., Parkinson’s disease alters multisensory perception: Insights from the Rubber Hand Illusion. Neuropsychologia 97, 38–45 (2017).

32. M. Honma, Y. Terao, Modulation of time in Parkinson’s disease: a review and perspective on cognitive rehabilitation. Frontiers in psychiatry 15, 1379496 (2024).

33. A. Conte, N. Khan, G. Defazio, J. C. Rothwell, A. Berardelli, Pathophysiology of somatosensory abnormalities in Parkinson disease. Nature Reviews Neurology 9, 687–697 (2013).

34. T. Hummel, G. Kobal, H. Gudziol, A. Mackay-Sim, Normative data for the “Sniffin’Sticks” including tests of odor identification, odor discrimination, and olfactory thresholds: an upgrade based on a group of more than 3,000 subjects. European Archives of Oto-Rhino-Laryngology 264, 237–243 (2007).

35. S. Trentin, B. S. Fraiman de Oliveira, Y. Ferreira Felloni Borges, C. R. de Mello Rieder, Systematic review and meta-analysis of Sniffin Sticks Test performance in Parkinson’s disease patients in different countries. European Archives of Oto-Rhino-Laryngology, 1–23 (2022).

36. S. S. Schiffman, Physicochemical Correlates of Olfactory Quality: A series of physicochemical variables are weighted mathematically to predict olfactory quality. Science 185, 112–117 (1974).

37. R. M. Khan et al., Predicting odor pleasantness from odorant structure: pleasantness as a reflection of the physical world. Journal of Neuroscience 27, 10015–10023 (2007).

38. K. Pospichalova, J. Vodicka, A. Kopal, New test of odor pleasantness in Parkinson’s disease. Functional Neurology 31, 149 (2016).

39. J. Hudry, S. Thobois, E. Broussolle, P. Adeleine, J.-P. Royet, Evidence for deficiencies in perceptual and semantic olfactory processes in Parkinson’s disease. Chemical senses 28, 537–543 (2003).

40. A. Mrochen et al., From sweet to sweat: hedonic olfactory range is impaired in Parkinson’s disease. Parkinsonism & related disorders 22, 9–14 (2016).

41. J. Snoek, H. Larochelle, R. P. Adams, Practical bayesian optimization of machine learning algorithms. Advances in neural information processing systems 25, (2012).

42. K. Snitz et al., Smellspace: an odor-based social network as a platform for collecting olfactory perceptual data. Chemical senses 44, 267–278 (2019).

43. T. Fawcett, in Proceedings 2001 IEEE international conference on data mining. (Ieee, 2001), pp. 131–138.

44. A. Arzi et al., Olfactory sniffing signals consciousness in unresponsive patients with brain injuries. Nature 581, 428–433 (2020).

45. J. L. Feldman, C. A. Del Negro, P. A. Gray, Understanding the rhythm of breathing: So near, yet so far. Annual Review of Physiology 75, 423–452 (2013).

46. N. Sobel, R. M. Khan, C. A. Hartley, E. V. Sullivan, J. D. Gabrieli, Sniffing longer rather than stronger to maintain olfactory detection threshold. Chemical senses 25, 1–8 (2000).

47. S. Džeroski, B. Ženko, Is Combining Classifiers with Stacking Better than Selecting the Best One? Machine Learning 54, 255–273 (2004).

48. B. Zadrozny, C. Elkan, in Proceedings of the eighth ACM SIGKDD international conference on Knowledge discovery and data mining. (2002), pp. 694–699.

49. T. Tat et al., Diagnosing Parkinson’s disease via behavioral biometrics of keystroke dynamics. Science Advances 11, eadt6631 (2025).

50. Y. Yeshurun, N. Sobel, An odor is not worth a thousand words: from multidimensional odors to unidimensional odor objects. Annual review of psychology 61, 219–241 (2010).

51. E. S. Lelo de Larrea-Mancera et al., A Characterization of Central Auditory Processing in Parkinson’s Disease. Journal of Parkinson’s Disease 14, 999–1013 (2024).

52. E. Uc et al., Visual dysfunction in Parkinson disease without dementia. Neurology 65, 1907–1913 (2005).

53. J. E. Duda, U. Shah, S. E. Arnold, V. M.-Y. Lee, J. Q. Trojanowski, The expression of α-, β-, and γ-synucleins in olfactory mucosa from patients with and without neurodegenerative diseases. Experimental neurology 160, 515–522 (1999).

54. L. Silveira-Moriyama et al., Regional differences in the severity of Lewy body pathology across the olfactory cortex. Neuroscience letters 453, 77–80 (2009).

55. Y. Masaoka, N. Yoshimura, M. Inoue, M. Kawamura, I. Homma, Impairment of odor recognition in Parkinson’s disease caused by weak activations of the orbitofrontal cortex. Neuroscience letters 412, 45–50 (2007).

56. K. S. Utz et al., A multisensory deficit in the perception of pleasantness in Parkinson’s disease. Journal of Parkinson’s Disease 11, 2035–2045 (2021).

57. M. Wachowiak, All in a sniff: olfaction as a model for active sensing. Neuron 71, 962–973 (2011).

58. D. Rojas-Líbano, L. M. Kay, Interplay between sniffing and odorant sorptive properties in the rat. Journal of Neuroscience 32, 15577–15589 (2012).

59. A. Arzi, L. Rozenkrantz, Y. Holtzman, L. Secundo, N. Sobel, Sniffing patterns uncover implicit memory for undetected odors. Current Biology 24, (2014).

60. A. Arzi et al., Humans can learn new information during sleep. Nature neuroscience 15, 1460–1465 (2012).

61. L. Rozenkrantz et al., A mechanistic link between olfaction and autism spectrum disorder. Current biology 25, 1904–1910 (2015).

62. N. Sobel et al., An impairment in sniffing contributes to the olfactory impairment in Parkinson’s disease. Proceedings of the National Academy of Sciences 98, 4154–4159 (2001).

63. R. Dubbioso, F. Manganelli, H. R. Siebner, V. Di Lazzaro, Fast intracortical sensory-motor integration: a window into the pathophysiology of Parkinson’s disease. Frontiers in human neuroscience 13, 111 (2019).

64. M. Andelman-Gur et al., Discriminating Parkinson’s disease patients from healthy controls using nasal respiratory airflow. Communications Medicine 4, 233 (2024).

65. A. B. Rosenfeldt, T. Dey, J. L. Alberts, Aerobic exercise preserves olfaction function in individuals with Parkinson’s disease. Parkinson’s disease 2016, 9725089 (2016).

66. M. Barbado et al., Volumetric changes in the anterior olfactory nucleus of the rat after neonatal olfactory deprivation. Experimental neurology 171, 379–390 (2001).

67. H. Braak et al., Staging of brain pathology related to sporadic Parkinson’s disease. Neurobiology of aging 24, 197–211 (2003).

68. F. Corotto, J. Henegar, J. Maruniak, Odor deprivation leads to reduced neurogenesis and reduced neuronal survival in the olfactory bulb of the adult mouse. Neuroscience 61, 739–744 (1994).

69. G. U. Höglinger et al., A new dopaminergic nigro-olfactory projection. Acta neuropathologica 130, 333–348 (2015).

70. J. J. Kim, S. Bandres-Ciga, K. Heilbron, C. Blauwendraat, A. J. Noyce, Bidirectional relationship between olfaction and Parkinson’s disease. npj Parkinson’s Disease 10, 1–8 (2024).

71. K. S. M. Taylor, J. A. Cook, C. E. Counsell, Heterogeneity in male to female risk for Parkinson’s disease. *Journal of Neurology*, Neurosurgery & Psychiatry 78, 905–906 (2007).

72. J. Wu et al., A nanoparticle-based wireless deep brain stimulation system that reverses Parkinson’s disease. Science Advances 11, eado4927 (2025).

73. N. Vijiaratnam, T. Simuni, O. Bandmann, H. R. Morris, T. Foltynie, Progress towards therapies for disease modification in Parkinson’s disease. The Lancet Neurology 20, 559–572 (2021).

74. N. D. Raig et al., Type II kinase inhibitors that target Parkinson’s disease–associated LRRK2. Science Advances 11, eadt2050 (2025).

75. G. Marino et al., Intensive exercise ameliorates motor and cognitive symptoms in experimental Parkinson’s disease restoring striatal synaptic plasticity. Science Advances 9, eadh1403 (2023).

76. R. Kahana-Zweig et al., Measuring and characterizing the human nasal cycle. PLoS ONE 11, e0162918–e0162918 (2016).

77. E. Erdfelder, F. Faul, A. Buchner, A. G. Lang, Statistical power analyses using G*Power 3.1: Tests for correlation and regression analyses. Behavior Research Methods 41, 1149–1160 (2009).

78. J. T. Giacino et al., Comprehensive systematic review update summary: disorders of consciousness: report of the guideline development, dissemination, and implementation subcommittee of the American Academy of Neurology; the American Congress of Rehabilitation Medicine; and the National Institute on Disability, Independent Living, and Rehabilitation Research. Neurology 91, 461–470 (2018).

