## Supplemental matrials for "The World Smells Different in Parkinson’s Disease"

### Supplementary Materials

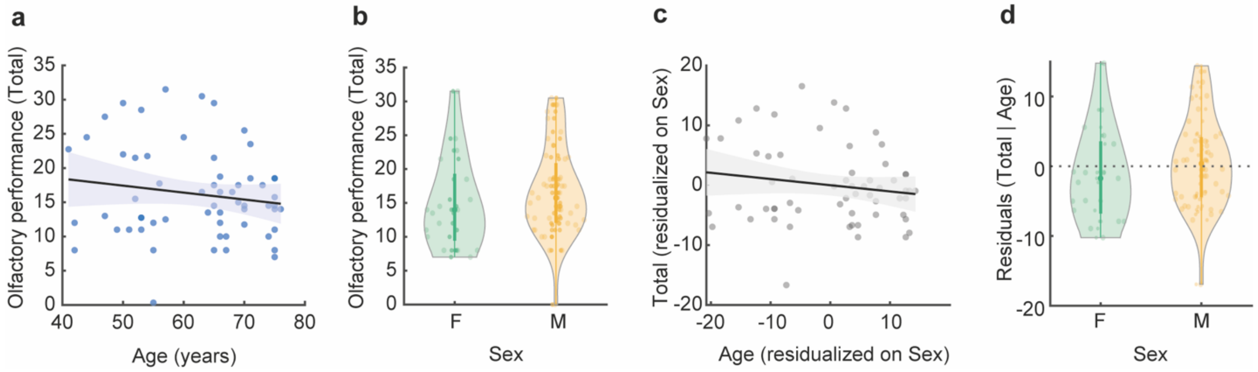


**Supp. Fig. 1. Sex- and age-related variation in olfactory performance (PD and OD only)**

**a,** Scatter of Sniffin' Sticks Total score vs. age (years) with a least-squares fit (black line) and 95% CI (light-blue band); each circle is one participant. **b,** Violin plots of Sniffin' Sticks Total score by sex (female = green, male = orange) with individual points overlaid. **c,** Added-variable (partial regression) plot showing the relation between model residuals of Sniffin' Sticks Total score (after removing sex) and age residualized on sex; black line represents the fit, grey band the 95% CI; each circle is one participant. **d,** Violin plots of age-residualized Total by sex (female = green, male = orange) with individual points overlaid.

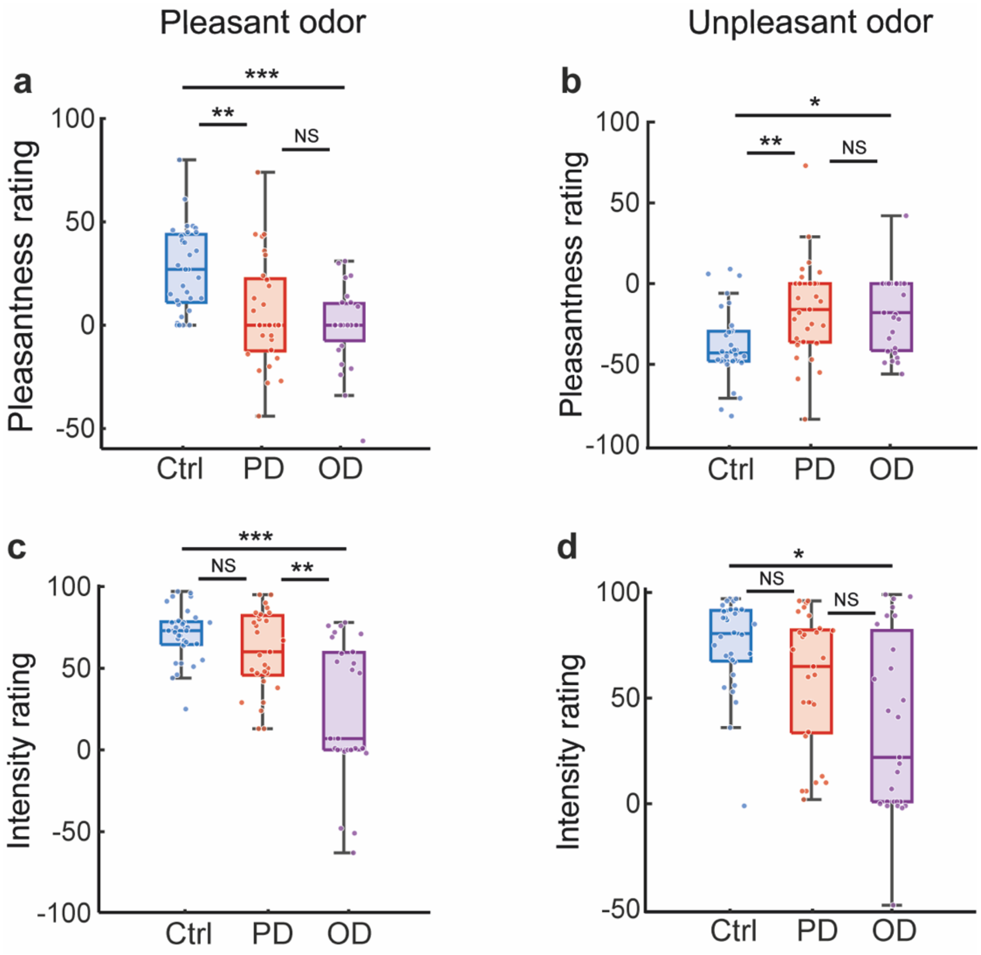


**Supp Fig. 2: Pleasantness ratings in PD and OD normalized by ratings of blank**

**a-d**, Box plots showing subjective ratings (VAS scale, 1-100), normalized by substruction to blank, of pleasantness (a,b) and intensity (c,d) for pleasant (a,c) and unpleasant (d,e) odorants in PD (red, n = 29), OD (purple, n = 27), and control (blue, n = 32) group. Central lines indicate median, top and bottom edges of the box indicate 75[th] and 25[th] percentiles, respectively. Whiskers reflect 1.5 times the interquartile range. Each circle represents a participant. *$P \leq 0.05$, **$P \leq 0.01$, ***$P \leq 0.001$.

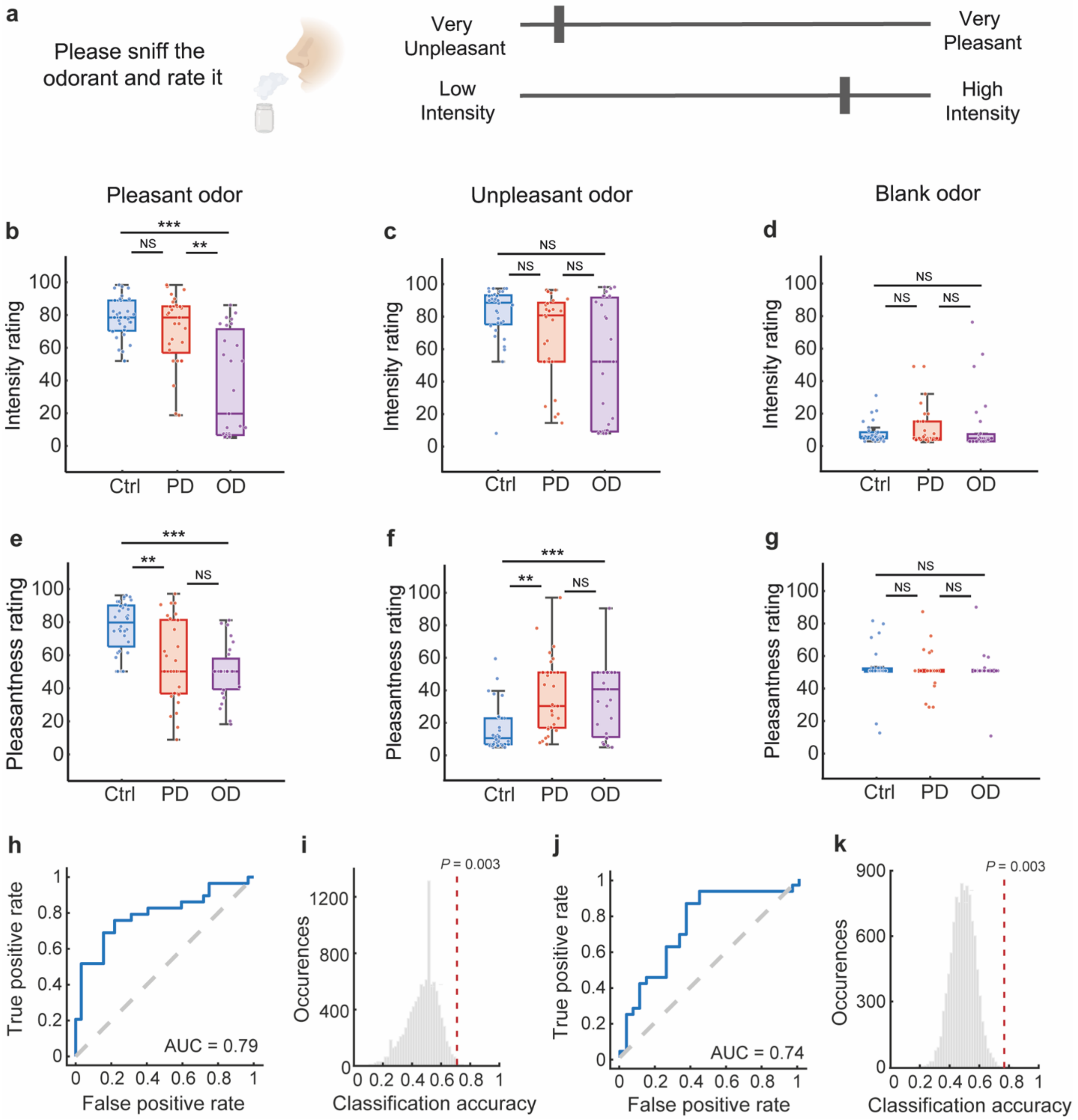


**Supp Fig. 3: Shifted primary axes of olfactory perception in Parkinson's disease**

**a,** Illustration of task procedure. **b-g**, Box plots displaying subjective ratings (VAS scale, 1-100) of intensity (b,c,d) and pleasantness (e,f,g) for pleasant (b,e), unpleasant (c,f), and blank (d,g) odorants in PD (red, n = 29), OD (purple, n = 27) and healthy controls (blue, n = 32). Central lines indicate median, top and bottom edges of the box indicate 75th and 25th percentiles, respectively. Whiskers reflect 1.5 times the interquartile range. Each circle represents a participant. *$P \leq 0.05$, **$P \leq 0.01$, ***$P \leq 0.001$. **h, j**, ROC curves for detecting PD from healthy controls (h) and for detecting PD from OD participants (j) using an SVM classifier on pleasantness and intensity ratings. *i, k*, Histogram of classification accuracy using an SVM classifier for shuffled labels data of PD and controls (i) and PD and OD (k) with actual classification accuracy marked by red dashed line. P-values are marked on top of the line.

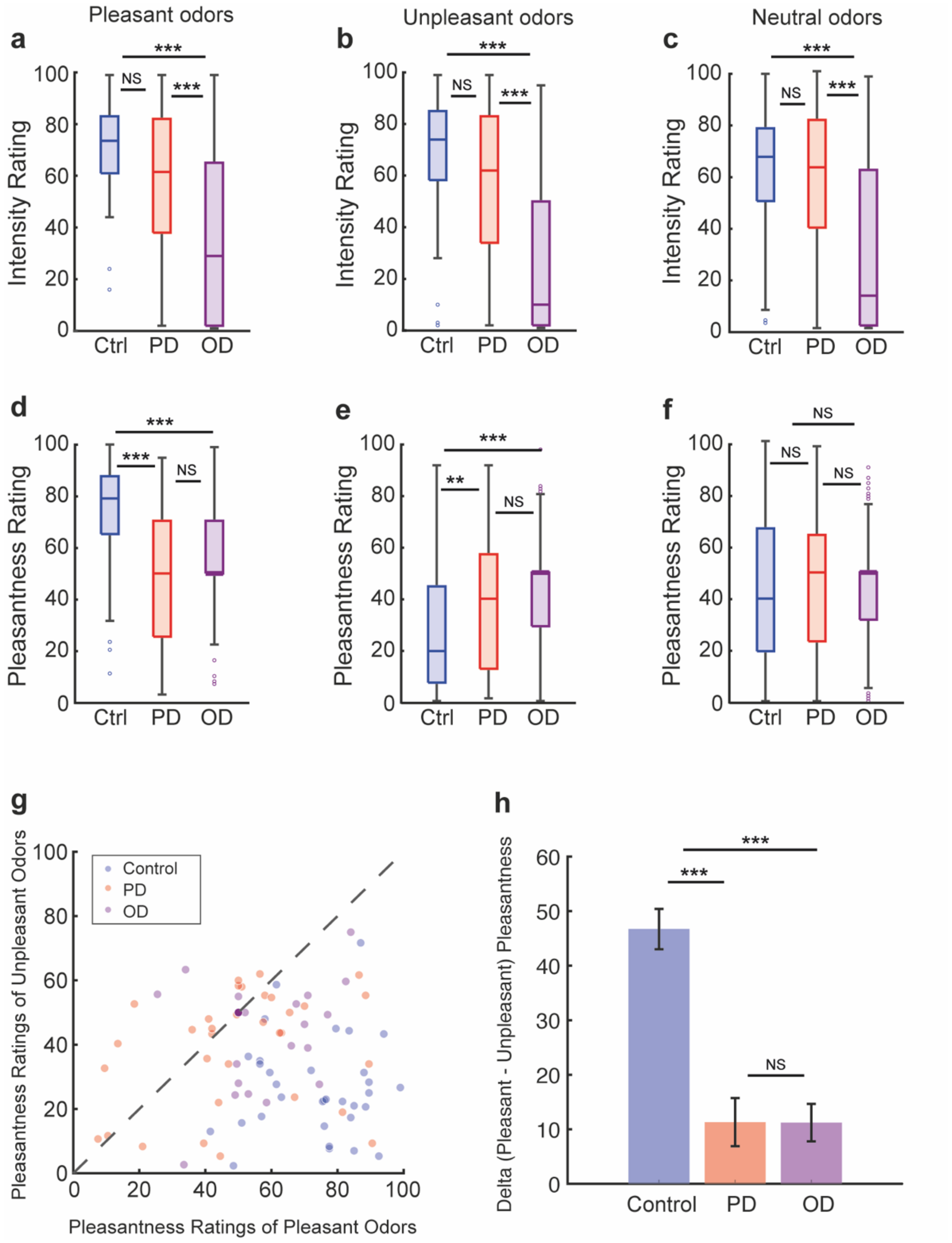

Pleasant odors
Unpleasant odors
Neutral odors
a
b
c
d
e
f
g
h
Intensity Rating
Pleasantness Rating
Ctrl
PD
OD
NS
***
**
Control
PD
OD
Pleasantness Ratings of Unpleasant Odors
Pleasantness Ratings of Pleasant Odors
Delta (Pleasant - Unpleasant) Pleasantness

**Supp Fig. 4: Intensity and pleasantness ratings for 10 odorants**

**a-f**, Box plots illustrating subjective ratings (VAS scale, 1-100) of intensity (a-c) and pleasantness (d-f) for pleasant (a,d), unpleasant (b,e), and blank (c,f) odorants in PD (red, n = 33), OD (purple, n = 28) and healthy controls (blue, n = 33) groups. Central lines indicate median, top and bottom edges of the box indicate 75$^{th}$ and 25$^{th}$ percentiles, respectively. Whiskers reflect 1.5 times the interquartile range. Each circle represents a participant. **g**, Scatter plot displaying pleasantness ratings of pleasant vs. unpleasant odorants in Control (blue), PD (red), and OD (purple) groups. Each circle represents a participant. h, Bar graph (mean +/- SEM) illustrating the difference (delta) between pleasantness ratings of pleasant and unpleasant odorants in each participant. *$P \leq 0.05$, **$P \leq 0.01$, ***$P \leq 0.001$.

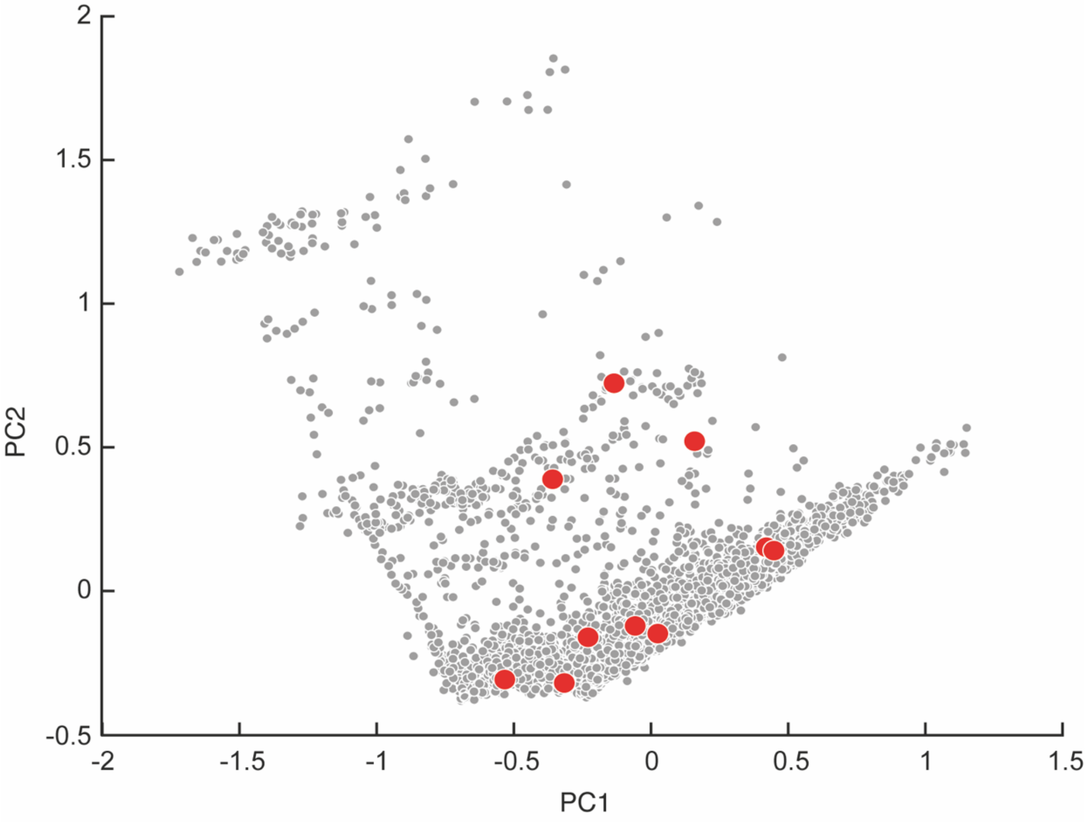


The 10

**Supp Fig. 4:**

**Supp Fig. 5: Selected monomolecules in physicochemical space**

monomolecules used in the study (red circles) depicted within the first and second principal components of a representative physicochemical space containing 4886 odorant molecules (grey circles). The 10 monomolecules are: Fenchone (CAS 7787-20-4), Isoamyl acetate (CAS 123-92-2), 3-Propylidene phthalide (CAS 17369-59-4), Cuminaldehyde (CAS 122-03-2), Strawberry glycidate 1 (CAS 77-83-8), Nonanal (CAS 124-19-6), Citral (CAS 5392-50-5), Skatole (CAS 83-34-1), Hexanol (CAS 111-27-3), 6-Methylquinoline (CAS 91-62-3). All monomolecules were rates across 11 descriptors: “Pleasantness”; “Intensity”; “Garlicky”; “Sweet”; “Fruity”; “Chemical”; “Bitter”; “Burnt”; “Spicy”; “Clean-Dirty”; “Fresh-Rotten.”

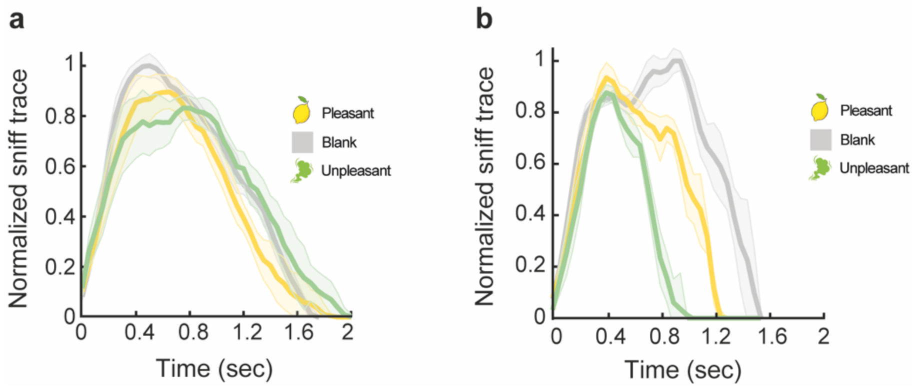


**Supp Fig. 6: Individual examples of sniff-response patterns**

**a**, Example traces of altered sniff response from participant with PD 009, sniff trace was normalized to the participant's maximal peak value. Normalized sniff duration (mean ± s.d.): pleasant (yellow), 0.92 ± 0.14 NDU (10 repetitions); unpleasant (green), 1.0 ± 0.14 NDU (9 repetitions); blank (grey), 1.0 ± 0.14 NDU (10 repetitions). **b**, Example traces of intact sniff response from healthy control 004, sniff trace was normalized to the participant's maximal peak value. Normalized sniff duration: pleasant (yellow), 0.87 ± 0.11 NDU (10 repetitions); unpleasant (green), 0.7 ± 0.08 NDU (10 repetitions); blank (grey), 1.06 ± 0.11 NDU (10 repetitions). Traces are shown as mean (line) and SEM (shaded areas).

**Supp Table 1: Clinical and demographic characteristics**

| Characteristic | | Individuals with PD (n = 33) | Control (n = 33) | OD patients (n = 28) |
|---|---|---|---|---|
| Age, mean (SD), years | | 65.8 (7.2) | 64.2 (7.5) | 58.1 (11.5) |
| Male sex, No. (%) | | 29 (88%) | 29 (88%) | 15 (54%) |
| Education, years | | 16.4 (2.7) | 17.2 (3.7) | 14.6 (3.1) |
| Right hand dominance, No. (%) | | 28 (85%) | 28 (85%) | 25 (89%) |
| MoCA score, mean (SD) | | 24.8 (3.9) | 25.7 (2.0) | 25.5 (3.4) |
| Beck score, mean (SD) | | 11.5 (8.2) | 4.9 (4.8) | 8.5 (9.1) |
| PD Disease duration, median (SD, range), years | | 6 (5.4, 1-29) | - | - |
| MDS-UPDRS Part III, mean (SD) | | 29.3 (13.3) | - | - |
| MDS-UPDRS total, mean (SD) | | 56.0 (22.0) | - | - |
| Hoehn and Yahr stage (median, SD) | | 2 (0.7) | - | - |
| Hoehn and Yahr Stage I, No. (%) | | 7 (21%) | - | - |
| Hoehn and Yahr Stage II, No. (%) | | 19 (58%) | - | - |
| Hoehn and Yahr Stage III, No. (%) | | 7 (21%) | - | - |
| Hoehn and Yahr Stage IV, No. (%) | | 0 (0%) | - | - |
| Medication for PD | Any, No. (%) | 30 (91%) | - | - |
| | Levodopa and other dopa derivatives, No. (%) | 22 (67%) | - | - |
| | Monoamine oxidase type B inhibitors, No. (%) | 10 (30%) | - | - |
| | Dopamine agonists, No. (%) | 19 (58%) | - | - |
| Acquired olfactory dysfunction cause | Upper respiratory tract infection (post-viral, inc. COVID-19), No. (%) | - | - | 9 (32%) |
| | Sinonasal disease, No. (%) | - | - | 8 (29%) |
| | Head trauma, No. (%) | - | - | 3 (11%) |
| | Unknown/Idiopathic, No. (%) | - | - | 8 (29%) |
